# Household solid fuel use and the risk of sarcopenia among middle-aged and older adults in China: the first nationwide cross-sectional and longitudinal study

**DOI:** 10.1101/2023.02.17.23286116

**Authors:** Wenming Shi, Tiantian Zhang, Yongsheng Yu, Li Luo

## Abstract

**Background:** Little is known about the association between household solid fuel use and sarcopenia risk. Using a nationally representative survey, we investigated the association between solid fuel used for heating and cooking and sarcopenia risk among middle-aged and older Chinese adults.

**Methods:** We leveraged two waves of data from the China Health and Retirement Longitudinal Study (CHARLS); 12,723 participants aged ≥ 45 years from 28 provinces across China were enrolled in 2011. Sarcopenia status was classified according to the Asian Working Group for Sarcopenia 2019 criteria. A total of 3,110 participants without sarcopenia were recruited at baseline and were followed up until 2013. Primary fuel types and durations used for heating and cooking were assessed using a validated questionnaire. Multinomial logistic regression explored the cross-sectional and longitudinal associations between solid fuel use and different stages of sarcopenia.

**Findings:** The prevalence of possible sarcopenia and sarcopenia were 34·8% and 5·7%, respectively. Compared with clean fuel users, those using solid fuel for both heating and cooking had significantly higher risks of possible sarcopenia (odds ratio, [OR] 1·72, 95%CI: 1·54-1·91) and sarcopenia (OR 1·74, 1·31-2·31). During the two-year follow-up, 569 participants (18·3%) developed with possible sarcopenia and 86 (2·8%) had sarcopenia. In the longitudinal analyses, solid fuel use was positively associated with the risk of possible sarcopenia (OR 1·75, 1·32-2·31), and the association was higher in individuals with a longer duration of solid fuel use. However, no significant association was observed with the incidence of sarcopenia. Older adults, with less physical activity might have a higher risk of sarcopenia when exposed to solid fuel.

**Interpretation:** Household solid fuel is associated with a higher risk of sarcopenia among middle-aged and older Chinese adults. These findings provide novel evidence for prioritizing public health policies to promote healthy aging by reducing solid fuel use.

**Funding:** National Natural Science Foundation of China

**Research in context:** *Evidence before this study:* We searched PubMed, Google Scholar, and the China National Knowledge Infrastructure for studies published in English and Chinese up to February 1, 2023. We used the search terms (“sarcopenia”) AND (“solid fuel” OR “biomass fuel” OR “coal” OR “air pollution”) and found a recent study that explored the cross-sectional relationship between ambient air pollution and sarcopenia risk in the UK. However, no large population-based study has investigated the effects of household solid fuel use on sarcopenia.

*Added value of this study:* Our study showed for the first time that household solid fuel use is associated with a higher risk of sarcopenia among middle-aged and older Chinese adults. Our findings provide important prospective evidence linking solid fuels to an increased risk of sarcopenia. Reducing solid fuel use can be beneficial in preventing sarcopenia and promoting healthy aging in older adults. To our knowledge, this study is the largest nationwide cross-sectional and longitudinal study to date investigating the association between household fuel types and duration of solid fuel use with different stages of sarcopenia.

*Implications of all the available evidence:* Our findings underscore the importance of improving access to clean fuels to reduce the risk of sarcopenia associated with cooking and heating fuel use. This study extends the knowledge that prioritizes public health policies to mitigate the adverse effects of solid fuel use on sarcopenia and provides implications for further mechanistic research.

## Introduction

Sarcopenia is a disease characterized by progressive loss of muscle mass and strength and is an important health concern facing aging societies.^1^ It is estimated that the prevalence of sarcopenia ranges from 9·9% to 40·4% among the elderly worldwide.^2^ Growing evidence indicates that sarcopenia can increase the risk of multiple adverse health outcomes including cardiovascular disease (CVD), cancer, frailty, and mortality. ^3-6^ However, the definition of sarcopenia is still developing as new findings challenge the current understanding, and many clinicians remain unaware of this condition.^4,7,8^ Given the rapid aging worldwide, identifying the potential risk factors for sarcopenia is promising for early prevention and reducing the burden of the disease.

Air pollution is recognized as the largest environmental risk factor for all-cause mortality globally. ^9^ In recent years, an increasing number of studies have shown that air pollution increases the risk of decreased handgrip strength, low skeletal muscle mass, or limited physical performance, which are components of sarcopenia.^10,11^ Though previous studies have examined select sarcopenia components, the lack of a standardized definition of sarcopenia poses uncertainty about the effects of air pollution exposure on sarcopenia.^12^ In 2019, the Asian Working Group for Sarcopenia (AWGS) defined sarcopenia by retaining the previous criteria of sarcopenia in AWGS 2014,^8^ and revising the diagnostic algorithm and cutoff values for each component.^13^ A recent cross-sectional study based on the UK Biobank database showed that exposure to ambient multiple air pollutants, such as fine particulate matter (PM_2.5_), PM_10_, and nitrogen oxide (NO_X_), increased the risk of sarcopenia.^12^ Nevertheless, long-term longitudinal studies are warranted to examine the effects of air pollution and help strengthen the causality.

Compared with the outdoor environment, it is suggested that the indoor environment is more closely related to people’s health.^14,15^ The elderly generally spend much of their time in a residential environment after retirement and may have longer exposure to their houses.^16^ As the main source of household air pollution (HAP), solid fuel use increases the risk of abundant adverse health effects, including cardiovascular disease (CVD), respiratory diseases, metabolic diseases, and all-cause mortality.^14,17^ Despite the many efforts to popularize the usage of clean fuel, approximately three billion people still rely on solid fuel (i.e., coal, crop residuals, wood, and dung) for cooking or other household needs, especially in developing countries including China.^18^ However, to our knowledge, no large-scale studies have been conducted to explore the association of household solid fuel use with sarcopenia risk in middle-aged and older adults.

To fill this research gap, we took advantage of a nationally representative survey in China, to carry out cross-sectional and longitudinal analyses to investigate the association of HAP from solid fuel use with different stages of sarcopenia among middle-aged and older adults. We aimed to provide scientific evidence regarding the etiology, early intervention, and prevention of sarcopenia.

## Methods

### Study design and participants

This study was based on the China Health and Retirement Longitudinal Study (CHARLS) project, a nationally representative survey of middle-aged and older adults in China. Details about the study design and sampling method have been described in previous literature.^19^ Briefly, the project used a multistage probability sampling strategy to collect high-quality data through one-on-one interviews using computer-assisted personal interviewing (CAPI) technology. The data included weights corrected for non-responses in individual and household samplings separately. The nationwide baseline survey was conducted in 2011, covering 450 villages and 150 counties, involved 17,708 respondents from 28 provinces across China. The CHARLS is an ongoing survey; the respondents were followed up every-two years, and a small set of new respondents were recruited during each survey. Information on the demographics, lifestyle and behaviour factors, physical measurements, and household environmental characteristics (i.e., area of residence, building structure) were recorded by trained staff. The CHARLS project was approved by the Ethical Review Committee of Peking University (# IRB00001052–11015). All participants provided written informed consent.

In our study, we extracted two waves of data (2011 and 2013) from the CHARLS. After excluding participants aged less than 45 years and those with missing data on sarcopenia status and household fuels use, 12,723 participants were screened in the cross-sectional analysis. We excluded 727 participants with sarcopenia and 4,425 participants with possible sarcopenia at baseline, resulting in 7,571 healthy participants for the follow-up survey. A total of 3,110 eligible individuals were included in the longitudinal analyses. Figure 1 shows the selection process of the participants in this study.

**Figure 1.**
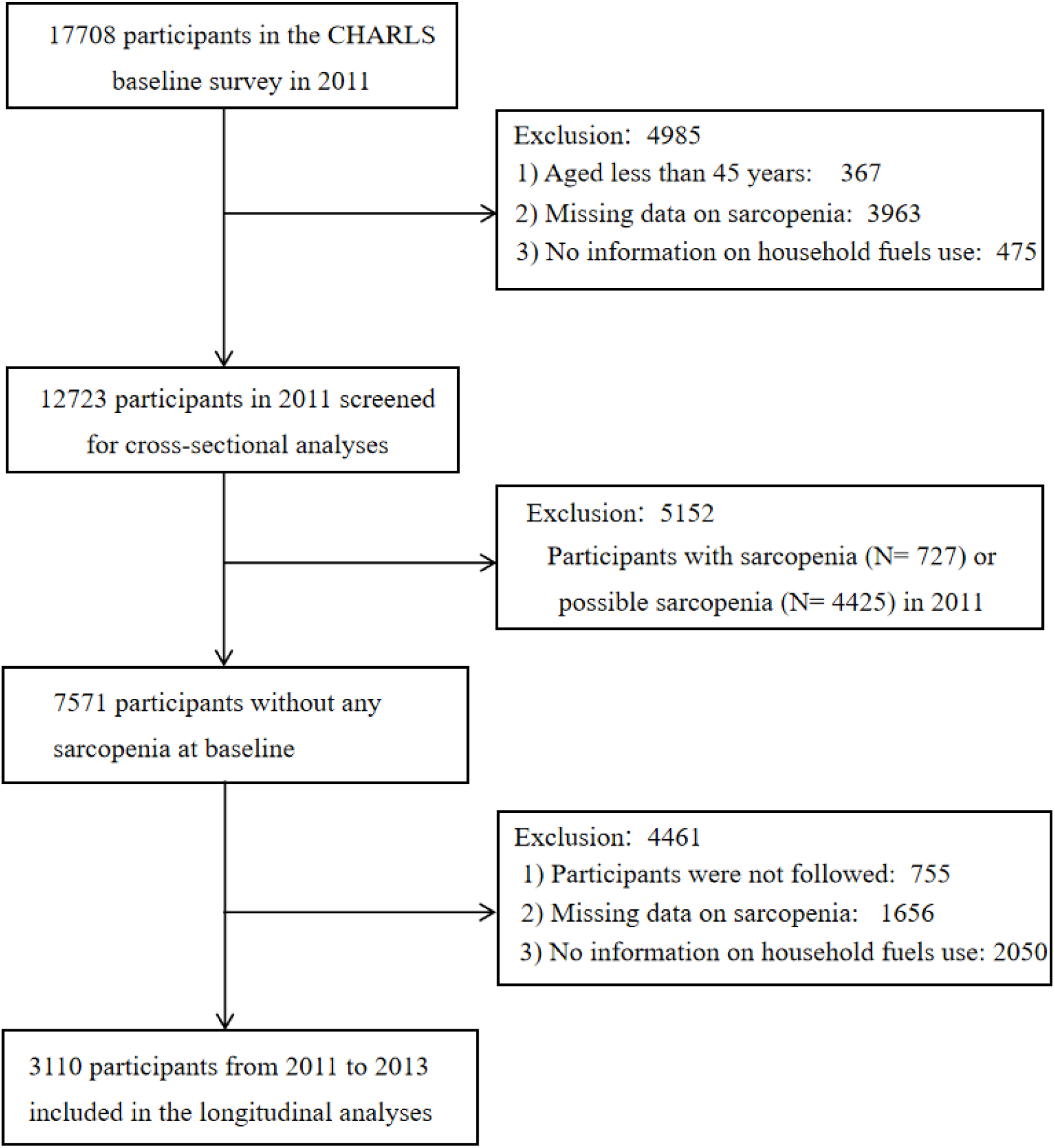
The flowchart of participants’ selection for cross-sectional and longitudinal analyses in the study.

### Assessment of sarcopenia status

Sarcopenia status was assessed based on the criteria of the AWGS 2019,^13^ which consists of three components: muscle strength, appendicular skeletal muscle mass (ASM), and physical performance. Sarcopenia is diagnosed when there is low muscle mass, low muscle strength or combined low physical performance. Possible sarcopenia was defined as low muscle strength with or without low physical performance.^20,21^

Handgrip strength (kg) was measured in the participants’ dominant hand and non-dominant hands by them squeezing a YuejianTM WL-1000 dynamometer as hard as possible. Each participant was tested in duplicates for both hands, and the average of the available maximum strength data was adopted. If one of the participants’ hands could not be measured for any reason, the maximum value of the other hand was used. The cut-off points for low muscle strength were < 28 kg and < 18 kg for men and women, respectively.^13^ The ASM was estimated by a validated formula for the Chinese population:^22^

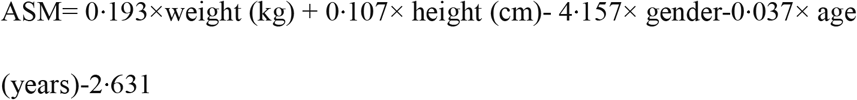

Several studies have shown that the ASM formula is in good agreement with dual-energy X-ray absorptiometry (DXA).^22,23^ The cut-off value for low muscle mass was based on the sex-specific lowest 20% of the height-adjusted muscle mass (ASM/ Height^2^) among the study participants,^21,23^ with < 7·01 kg/m^2^ in men and < 5·26 kg/m^2^ in women. For physical performance, gait speed and the five-time chair stand test were performed using the method described in previous literature,^21^ and low physical performance was assessed based on the AWGS 2019 criteria.^13^

### Household air pollution exposure

HAP was evaluated using a surrogate of solid fuel used for heating and cooking. A validated CHARLS questionnaire was used to collect information about the main heating energy source and cooking fuel sources in their houses in each study wave. Solid fuel was defined based on the primary use of coal, crop residue, or wood burning. Those using solar, natural gas, liquefied petroleum gas, marsh gas, electricity, or others were categorized as clean fuels because they produced much less air pollution.^14,24^ The duration of solid fuel use was defined according to the type of fuel used in two waves, marked as three subgroups: less than one year, one to two years, and three years or more.

### Covariates

Information on socio-demographic characteristics, including gender, age, educational level, marital status, family’s economic status (≥ average level or < average level), and residence (urban or rural), was collected using a validated questionnaire during the CHARLS survey. Height and weight were measured and body mass index (BMI) was calculated as weight in kilograms divided by height in meters squared. Behaviour factors, including cigarette smoking status (current; former; or never) and alcohol consumption were also recorded. Similar to the method in the previous literature,^25^ physical activity (PA) levels were assessed in the CHARLS and classified into low, moderate, high, and unclear subgroups in this study.

### Statistical analysis

The descriptive statistics of the baseline characteristics of the participants according to the sarcopenia status was summarized and compared using the chi-square test, analysis of variance, or Kruskal-Wallis test. We also compared the baseline characteristics of the cross-sectional and longitudinal analytic participants according to the household fuel types for heating and cooking. The percentages of the various fuel types for heating and cooking were calculated separately at baseline and during the follow-up year. The prevalence and incidence of possible sarcopenia, sarcopenia, and sarcopenia components were also calculated.

Multinomial logistic regression was conducted to explore the cross-sectional associations between solid fuel use for heating and cooking (reference: clean fuel), separately or simultaneously, and the risk of different stages of sarcopenia. The longitudinal analyses on the different fuel types and duration of solid fuel use, as well as the incidence risk of possible sarcopenia and sarcopenia were also examined by logistic regression models.^20^ We performed three different models: Model 1: crude model; Model 2: adjusted for age, gender, BMI, marriage status, educational level, and the family’s economic status; Model 3: further adjusted for cigarette smoking, alcohol consumption, physical activity, and residence based on Model 2. Unclear answer in PA levels was grouped into a single category and kept in the regression model.^26^

Stratified analyses were performed to examine several potential modifiers of gender (male or female), age (≥ 60 years or <60 years), behavioral factors including smoking exposure (yes or no), alcohol consumption (yes or no), and PA level (low/ moderate or high; considering the low ratio of PA level, we merged individuals with low or moderate PA into a category). Moreover, we also investigated whether the building’s environmental characteristics, such as the area of the house, type of building (one-story or multi-story building), and structure of the building (concrete and steel; bricks and wood, mixed structure and others) influenced the associations.

Several sensitivity analyses were performed: 1) we investigated the association of solid fuel use for both heating and cooking at baseline with the incident risk of possible sarcopenia and sarcopenia; 2) we examined the cross-sectional and longitudinal associations of solid fuel use with sarcopenia components separately; and we tested the robustness of the association by performing cross-sectional and longitudinal analyses in a sub-sample with a clear classification of PA in the study.

All statistical analyses were conducted using STATA 16·0 (Stata Corp., College Station, TX, USA), and the significance level was set at *P* < 0·05.

## Results

A total of 12,723 participants were included in this cross-sectional analysis. Table 1 presents the baseline characteristics of the participants according to their sarcopenia status. The prevalence of possible sarcopenia and sarcopenia were 34·8% and 5·7%, respectively. Compared to healthy participants, individuals with sarcopenia are more likely to be older, have lower education, be unmarried or widowed, reside in rural areas, have poorer financial status, and use solid fuels at home. Table S1 shows the baseline characteristics of the participants included in both the cross-sectional and longitudinal analyses, across the categories of household fuel types used for heating and cooking. We observed a very similar distribution of the participants’ characteristics, except for a minor difference in gender (Table S1).

**Table 1.** Baseline characteristics of the participants according to sarcopenia status ^#^.

| Characteristics | No sarcopenia<br>(N= 7571) | Possible sarcopenia<br>(N= 4425) | Sarcopenia<br>(N= 727) | P-value |
| --- | --- | --- | --- | --- |
| Age (years) | 56.9 ± 8.2 | 61.5 ± 9.6 | 72.1 ± 9.2 | < 0.001 |
| Gender |  |  |  |  |
| Male | 3913 (51.7) | 1823 (41.2) | 335 (46.1) | < 0.001 |
| Female | 3658 (48.3) | 2602 (58.8) | 392 (53.9) |  |
| BMI (kg/m<sup>2</sup>) | 23.57 ± 3.49 | 23.75 ± 3.65 | 18.97 ± 2.02 | < 0.001 |
| Educational level |  |  |  |  |
| ≤ Primary school | 4681 (61.8) | 3400 (76.8) | 669 (92.0) | < 0.001 |
| Middle school | 1846 (24.4) | 717 (16.2) | 36 (5.0) |  |
| ≥ High school | 1044 (13.8) | 308 (7.0) | 22 (3.0) |  |
| Marital status |  |  |  |  |
| Married | 6882 (90.9) | 3714 (83.9) | 465 (64.0) | < 0.001 |
| Unmarried or widow | 689 (9.1) | 711 (16.1) | 262 (36.0) |  |
| Residence |  |  |  |  |
| Urban | 2954 (39.0) | 1499 (33.9) | 187 (25.7) | < 0.001 |
| Rural | 4617 (61.0) | 2926 (66.1) | 540 (74.3) |  |
| Family's economic status |  |  |  |  |
| ≥ Average level | 4221 (55.8) | 2361 (53.4) | 344 (47.3) | 0.002 |
| < Average level | 3253 (43.0) | 1969 (44.5) | 343 (47.2) |  |
| Physical activity |  |  |  |  |
| Low | 592 (7.8) | 472 (10.7) | 61 (8.4) | < 0.001 |
| Moderate | 537 (7.1) | 341 (7.7) | 50 (6.9) |  |
| High | 2037 (26.9) | 1106 (25.0) | 166 (22.8) |  |
| Unclear | 4405 (58•2) | 2506 (56•6) | 450 (61•9) |  |
| Cigarette smoking |  |  |  |  |
| Current | 2522 (33•3) | 1223 (27•6) | 235 (32•3) |  |
| Former | 660 (8•7) | 383 (8•7) | 67 (9•2) | <0•001 |
| Never | 4389 (58•0) | 2819 (63•7) | 425 (58•5) |  |
| Alcohol consumption |  |  |  |  |
| Yes | 3374 (44•6) | 1603 (36•2) | 269 (37•0) |  |
| No | 4197 (55•4) | 2822 (63•8) | 458 (63•0) | <0•001 |
| Type of household fuels |  |  |  |  |
| Clean fuels for heating & cooking | 2605 (34•4) | 1056 (23•9) | 123 (16•9) |  |
| Mixed fuels for heating & cooking | 2140 (28•3) | 1236 (27•9) | 204 (28•1) | < 0•001 |
| Solid fuels for heating & cooking | 2826 (37•3) | 2133 (48•2) | 400 (55•0) |  |
| Handgrip strength (kg) | 34•53 ± 12•33 | 27•68 ± 12•10 | 17•48 ± 5•85 | <0•001 |
| ASM/ Ht<sup>2</sup>, (kg/m<sup>2</sup>) | 6•89 ± 1•07 | 6•65 ± 1•07 | 5•53 ± 1•01 | < 0•001 |

The percentages of different fuel types used for heating and cooking at baseline and follow-up are presented in Figure S1. Overall, the percentages of solid fuel use decreased for both heating (from 55·9% to 48·3%) and cooking (from 56·5% to 45·4%) from 2011 to 2013. Figure S2 shows the prevalence and incidence of sarcopenia components across the different fuel types. Generally, individuals who use solid fuels have a higher prevalence and incidence of the components of sarcopenia (Figure S2).

Table 2 shows the cross-sectional associations between different fuel types used for heating and cooking and the risk of possible sarcopenia and sarcopenia in 2011. Compared with clean fuel use for household heating, solid fuel use was positively associated with a higher risk of possible sarcopenia (adjusted odds ratio, aOR 1·58, 95% CI: 1·45-1·73, Model 3) and sarcopenia (aOR 1·44, 1·17-1·79, Model 3) separately, after controlling for confounders (Table 2). For the specific type of solid fuels for heating, the estimates for the association of participants who used coal, or crop residue/ wood were 1·44 (1·30-1·60), 1·71 (1·55-1·89) for possible sarcopenia, and 1·30 (1·03-1·66), 1·63 (1·26-2·09) for sarcopenia, respectively. Similar associations were also observed for household cooking (Table 2). Compared with clean fuel for both heating and cooking, solid fuel users had a higher risk of possible sarcopenia (1·72, 1·54-1·91) and sarcopenia (1·74, 1·31-2·31), respectively (Table 2).

**Table 2.**
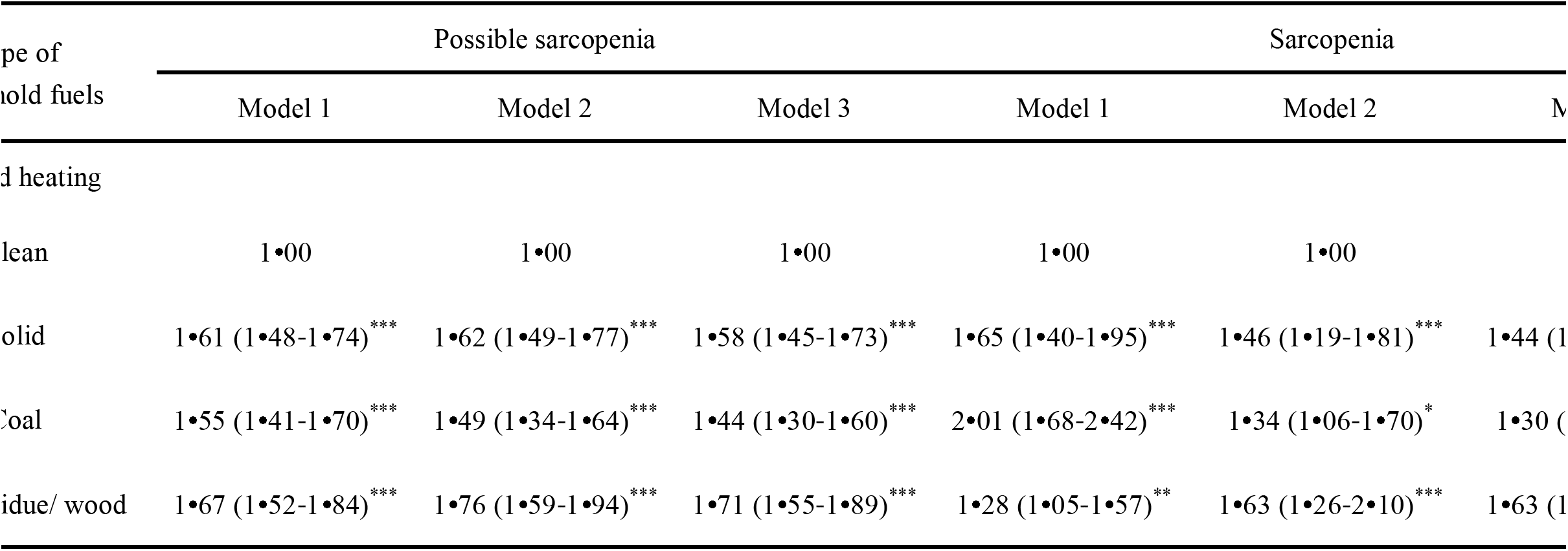
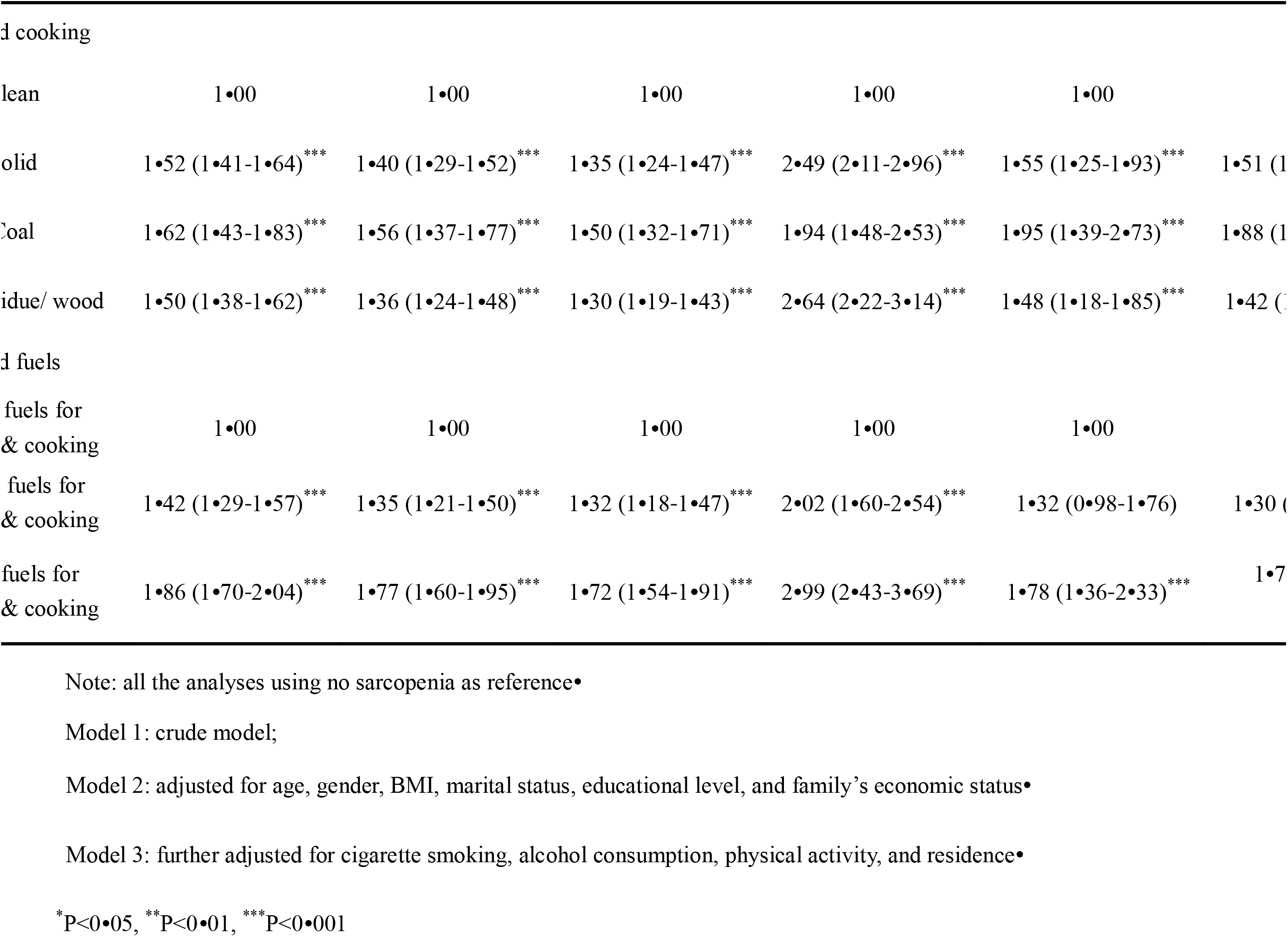
Cross-sectional associations between household polluting fuel use and different stages of sarcopenia.

Of the 3,110 longitudinal analytic samples, 569 (18·3%) and 86 (2·8%) participants developed possible sarcopenia and sarcopenia, respectively, from 2011 to 2013. Table3 shows the incidence of possible sarcopenia and sarcopenia across various household heating and cooking fuel types. In the crude model, solid fuel use was significantly associated with a higher risk of incident possible sarcopenia and sarcopenia than clean fuel use. After controlling for confounders, significant associations were also observed in the possible sarcopenia groups (1·75, 1·32-2·31), but not for sarcopenia (Table 3). In the sensitivity analyses, we found a robust association between solid fuel use at baseline and the incident risk of possible sarcopenia (Table S2). Table 4 presents the longitudinal associations between the duration of solid fuel use for heating and cooking separately, and the risk of possible sarcopenia and sarcopenia. We found that a longer duration of solid fuel use, both in heating and cooking, was positively associated with a higher risk of possible sarcopenia (Table 4). However, no significant association was observed for the incidence of sarcopenia (Table 4).

**Table 3.** Longitudinal associations between household polluting fuels use and different stages of sarcopenia.

| Outcomes | Clean fuels for<br>heating & cooking<br>(N= 837) | Mixed fuels for<br>heating & cooking<br>(N= 1519) | Solid fuels for<br>heating & cooking<br>(N= 754) |
| --- | --- | --- | --- |
| Possible sarcopenia |  |  |  |
| Cases (%) | 127 (15•2) | 246 (16•2) | 196 (26•0) |
| Model 1 | 1•00 (Ref) | 1•09 (0•86-1•38) | 2•02 (1•57-2•60)*** |
| Model 2 | 1•00 (Ref) | 1•02 (0•80-1•31) | 1•86 (1•43-2•42)*** |
| Model 3 | 1•00 (Ref) | 0•97 (0•75-1•25) | 1•75 (1•32-2•31)*** |
| Sarcopenia |  |  |  |
| Cases (%) | 16 (1•9) | 42 (2•8) | 28 (3•7) |
| Model 1 | 1•00 (Ref) | 1•48 (0•83-2•65) | 2•29 (1•23-4•28)** |
| Model 2 | 1•00 (Ref) | 1•04 (0•54-2•02) | 1•35 (0•65-2•79) |
| Model 3 | 1•00 (Ref) | 0•93 (0•46-1•88) | 1•16 (0•53-2•54) |

Note: all the analyses using no sarcopenia as reference•
Model 1: crude model;
Model 2: adjusted for age, gender, BMI, marital status, educational level, and family's economic status•
Model 3: further adjusted for cigarette smoking, alcohol consumption, physical activity, and residence•
\*\*P<0•01, \*\*\*P<0•001

**Table 4.** Longitudinal associations between duration of solid fuels use and different stages of sarcopenia.

| Duration of solid<br>fuels use | Possible sarcopenia |  |  | Sarcopenia |  |  |
| --- | --- | --- | --- | --- | --- | --- |
|  | Model 1 | Model 2 | Model 3 | Model 1 | Model 2 | Model 3 |
| Household heating |  |  |  |  |  |  |
| Less than one year | 1•00 | 1•00 | 1•00 | 1•00 | 1•00 | 1•00 |
| One to two years | 1•22<br>(0•95-1•56) | 1•18<br>(0•92-1•53) | 1•14<br>(0•88-1•48) | 2•14<br>(1•20-3•84)* | 1•55<br>(0•81-2•96) | 1•45<br>(0•75-2•82) |
| Three years or more | 1•77<br>(1•43-2•20) | 1•74<br>(1•40-2•18)*** | 1•68<br>(1•33-2•11)*** | 2•12<br>(1•22-3•67)** | 1•67<br>(0•90-3•09) | 1•56<br>(0•82-2•98) |
| Household cooking |  |  |  |  |  |  |
| Less than one year | 1•00 | 1•00 | 1•00 | 1•00 | 1•00 | 1•00 |
| One to two years | 1•21<br>(0•94-1•56) | 1•16<br>(0•90-1•50) | 1•11<br>(0•85-1•45) | 1•46<br>(0•78-2•72) | 0•87<br>(0•43-1•76) | 0•79<br>(0•38-1•63) |
| Three years or more | 1•67<br>(1•36-2•06)*** | 1•53<br>(1•23-1•90)*** | 1•45<br>(1•15-1•84)** | 2•27<br>(1•36-3•78)** | 1•36<br>(0•76-2•44) | 1•21<br>(0•64-2•28) |

Note: all the analyses using no sarcopenia as reference•
Model 1: crude model;
Model 2: adjusted for age, gender, BMI, marital status, educational level, and family's economic status•
Model 3: further adjusted for cigarette smoking, alcohol consumption, physical activity, and residence•
\* $P<0.05$ , \*\* $P<0.01$ , \*\*\* $P<0.001$

Stratified analyses showed that a significant association of solid fuel with sarcopenia was only observed in participants aged 60 years and older or in those with less PA (Figure 2). In the subgroup analyses by environmental characteristics, a significant association was only observed in solid fuel users living in smaller houses (Figure S3). However, there was no statistical difference in the association in other stratifications by building characteristics. In the sensitivity analyses of sarcopenia components, we observed inverse associations between solid fuel use and muscle strength and physical performance in both the cross-sectional and longitudinal analytic samples (Table S3, S4). Moreover, the sensitivity analyses showed very similar associations with different stages of sarcopenia in the sub-sample with clear PA information (Table S5).

**Figure 2.**
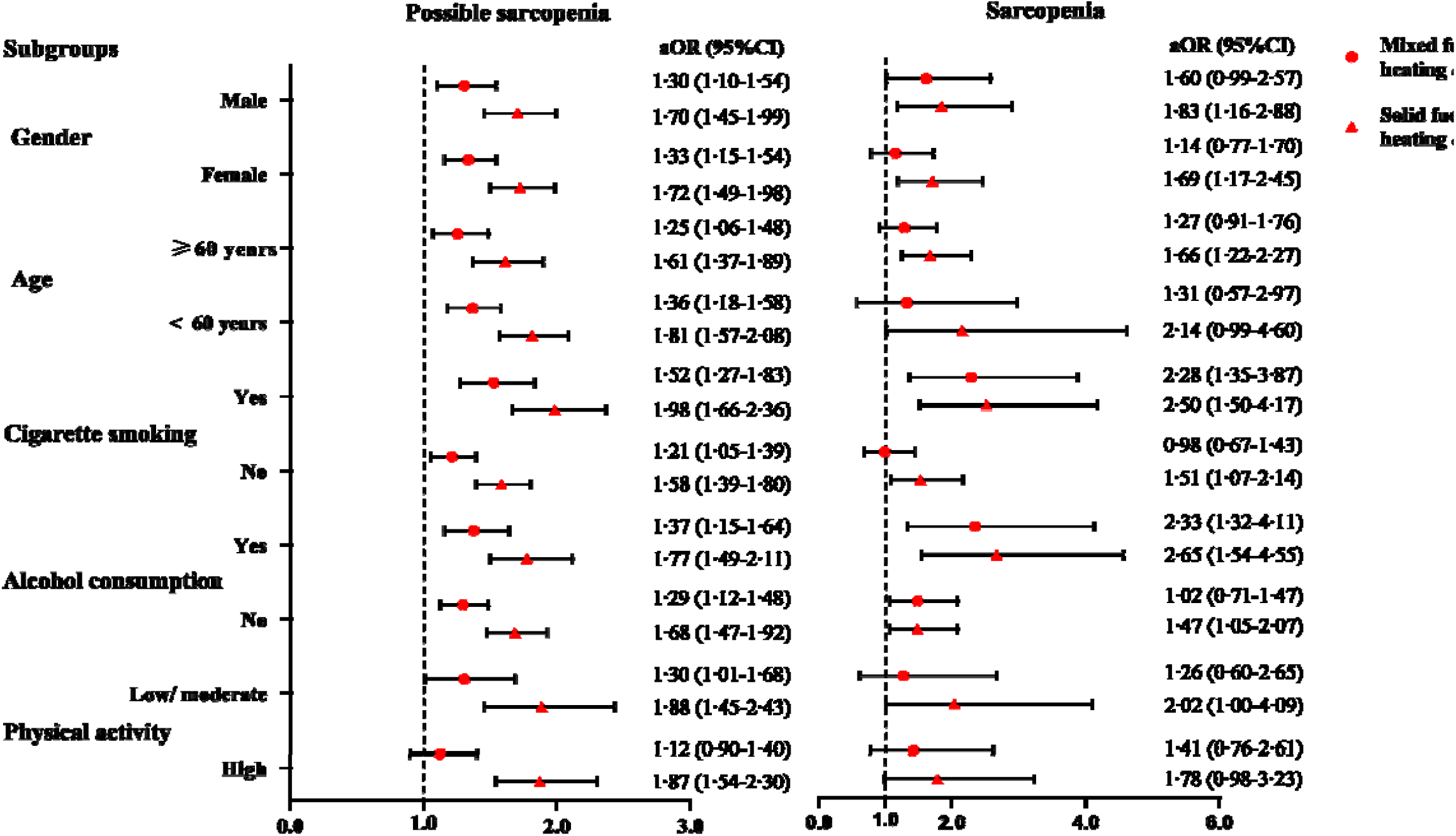
Cross-sectional associations between household solid fuels use and possible sarcopenia and sarcopenia stratified by gender, age and behavior factors^#^ ^#^adjusted for age, gender, BMI, marital status, educational level, family’s economic status, cigarette smoking, alcohol consumption, physical activity, and residence except for the stratified variable in the model•

## Discussion

This study is the first to investigate the association between household solid fuel use and the risk of different stages of sarcopenia among middle-aged and older adults. In the cross-sectional analyses, we found that, compared with clean fuel, use of solid fuel significantly increased the risk of both possible sarcopenia and sarcopenia. Participants aged 60 years and older or those with less PA levels might have a high risk of sarcopenia. In the longitudinal analyses, we observed that solid fuel use was significantly associated with the incidence risk of possible sarcopenia but not with incident sarcopenia. The robustness of the associations was demonstrated using sensitivity analysis.

In general, previous investigations on environmental factors have mostly focused on ambient air pollution, while no studies have explored the association between household solid fuel use and the risk of sarcopenia. A cross-sectional study based on the UK biobank database showed that long-term exposure to ambient air pollutants such as particulate matter and nitrogen dioxide increased the risk of probable sarcopenia.^12^ Compared with outdoor air pollution, the indoor environment is more closely related to people’s health.^14,15^ It is estimated that approximately 450 million people use solid fuels in China, ^27^ resulting in numerous toxic air pollutants and high concentrations of HAP.^28,29^ In our study, the association of household solid fuel use with different stages of sarcopenia was examined by cross-sectional and longitudinal analyses. These findings are similar to those of previous studies that investigated the effects of air pollution on different components of sarcopenia.^11,30,31^ However, previous studies have explored the selection of sarcopenia components without a standardized definition of sarcopenia, which might pose an uncertainty in the effects of air pollution on sarcopenia. Using the AWGS 2019 criteria, this study provides new evidence for household solid fuel use at different stages of sarcopenia.

However, the underlying mechanisms of this association remain unclear. There are several possible explanations. First, insufficient combustion of solid fuels can release multiple harmful substances, including particulate matter (PM), black carbon, nitrogen oxides, carbon monoxide, volatile organic compounds, and heavy metals.^28^ These chemicals easily to concentrate and reach the trachea, bronchi, and lungs through inhalation, and can further penetrate the alveoli into the blood circulation.^28^ Animal experiments showed that PM_2.5_ can induce oxidative stress and an inflammatory response, which may increase the loss of muscle strength and mass by causing oxidative damage to biomolecules.^32^ Moreover, PM and ozone exposure can increase mitochondrial oxidant production and mtDNA damage.^33^ It is reported that aging mitochondria may be another possible cause of sarcopenia, which is related to a loss of muscle fiber cross-sectional area.^34^

In addition to sarcopenia, the sensitivity analysis also showed a robust association between solid fuel use with certain sarcopenia components. In the stratified analysis, older adults, or those with less PA might have a higher risk of sarcopenia among solid fuel users. The reason for this may be that the risk of sarcopenia increases with age.^7^ It is suggested that skeletal muscle mass and function decline with aging, which restrains posture maintenance, balance, and quality of life in older adults.^35^ Moreover, lack of exercise has been a widely accepted risk factor for sarcopenia,^36^ similar to our results. In addition, it is well known that HAP is related to residential environments including household area and layout.^37^ In this study, we explored the influence of building environmental characteristics on the association of solid fuel users with sarcopenia. The results indicated that solid fuels users living in smaller houses had an increased risk of sarcopenia. This can be explained by the fact that the small size of a house may not be conducive to ventilation, which can affect the indoor pollutant levels.

Our study has several strengths. First, to the best of our knowledge, this is the first study to explore the association of household solid fuel use and possible sarcopenia and sarcopenia based on cross-sectional and longitudinal analyses in China. Our findings highlight that household solid fuel exposure, such as coal, crop residue, or wood, might be an important risk factor for sarcopenia. Second, this study included a large-scale, nationally representative sample, and the extrapolation of the results was of relatively high quality. The robustness of the association between solid fuel use and the risk of possible sarcopenia and sarcopenia has implications for developing public health policies aimed at reducing indoor air pollution. Third, our study added new evidence by uncovering the association between HAP and the risk of sarcopenia using the screening criteria developed by the AWGS. Fourth, we explored whether age, gender, and behavioral factors modified the associations of household solid fuel use with sarcopenia outcomes, something that has been under-examined. Finally, we evaluated the possible influence on individuals living in various residential building environments. These findings suggest that optimizing the structure of a building and increasing the clean fuel supply could help reduce the risk of sarcopenia.

This study has several limitations. First, we used proxy variables for HAP and did not directly monitor personal exposure to air pollutants, because such information could not be obtained in such a large-scale study. Second, owing to the observational design, it is difficult to establish a causal relationship between household solid fuel exposure and sarcopenia. Third, given the differences in culture and study populations, our findings might not be generalizable to younger age groups or populations in other countries. Fourth, considering the relatively high percentage of individuals with unclear information on their PA levels, this might restrict power in the stratified analysis. In the sensitivity analysis of a sub-sample with clear PA information, we observed very similar associations. Finally, despite a series of covariates being adjusted for in the study, unobserved or residual confounding factors, such as indoor ventilation habits and dietary factors, were not controlled in the model due to unavailable data.

## Conclusions

Using a nationally representative survey, our study provides new evidence on the association between household solid fuel use and different stages of sarcopenia among middle-aged and older Chinese adults. Longer durations of solid fuel use, and specific fuel types such as coal, crop residue, or wood burning are associated with a higher incident risk of possible sarcopenia. Older adults with less PA or living in small houses may have an increased risk of sarcopenia when exposed to solid fuels. Our findings highlight the prioritization of public health policies and interventions to mitigate the adverse effects of HAP by reducing solid fuel use and promoting healthy aging in China.

## Data Availability

All data produced in the present study are available upon reasonable request to the authors

## Contributors

WS and LL has the idea for the study and contributed to study design. WS and TZ analysed and interpreted data. WS wrote the draft, and TZ, YY, LL contributed to the critical revision of the manuscript. WS and LL supervised overall study implementation and obtained funding.

## Acknowledgement

We thank the Peking National Center for Economic Research for providing the CHARLS data. We are grateful to all the participants and researchers in this project.

---

**Figure S1.**
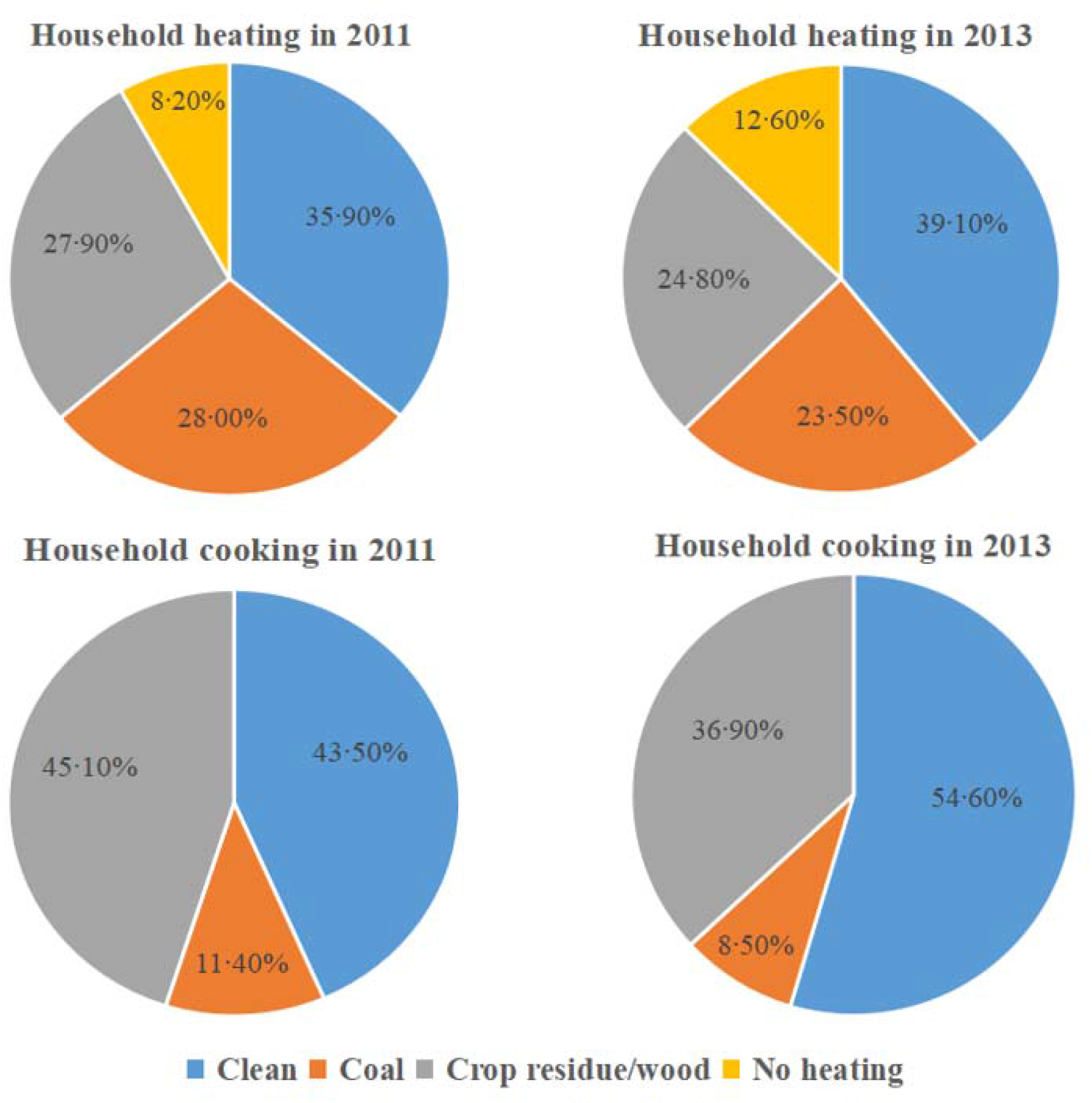
The percentages of different household fuels for heating or cooking in baseline and follow-up year.

**Figure S2.**
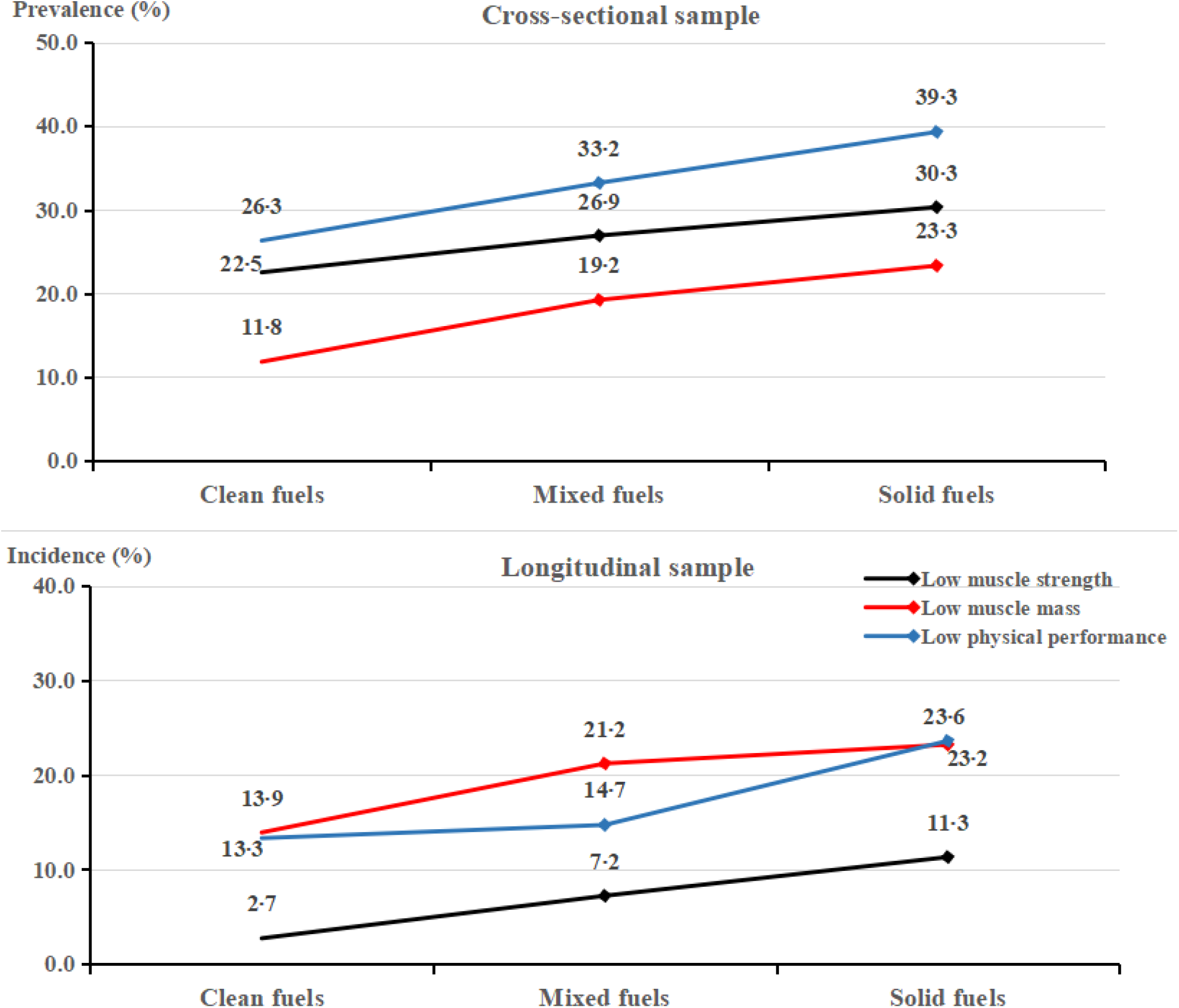
• The prevalence or incidence of sarcopenia components according to different fuels use for heating and cooking in baseline and follow-up year •

**Figure S3.**
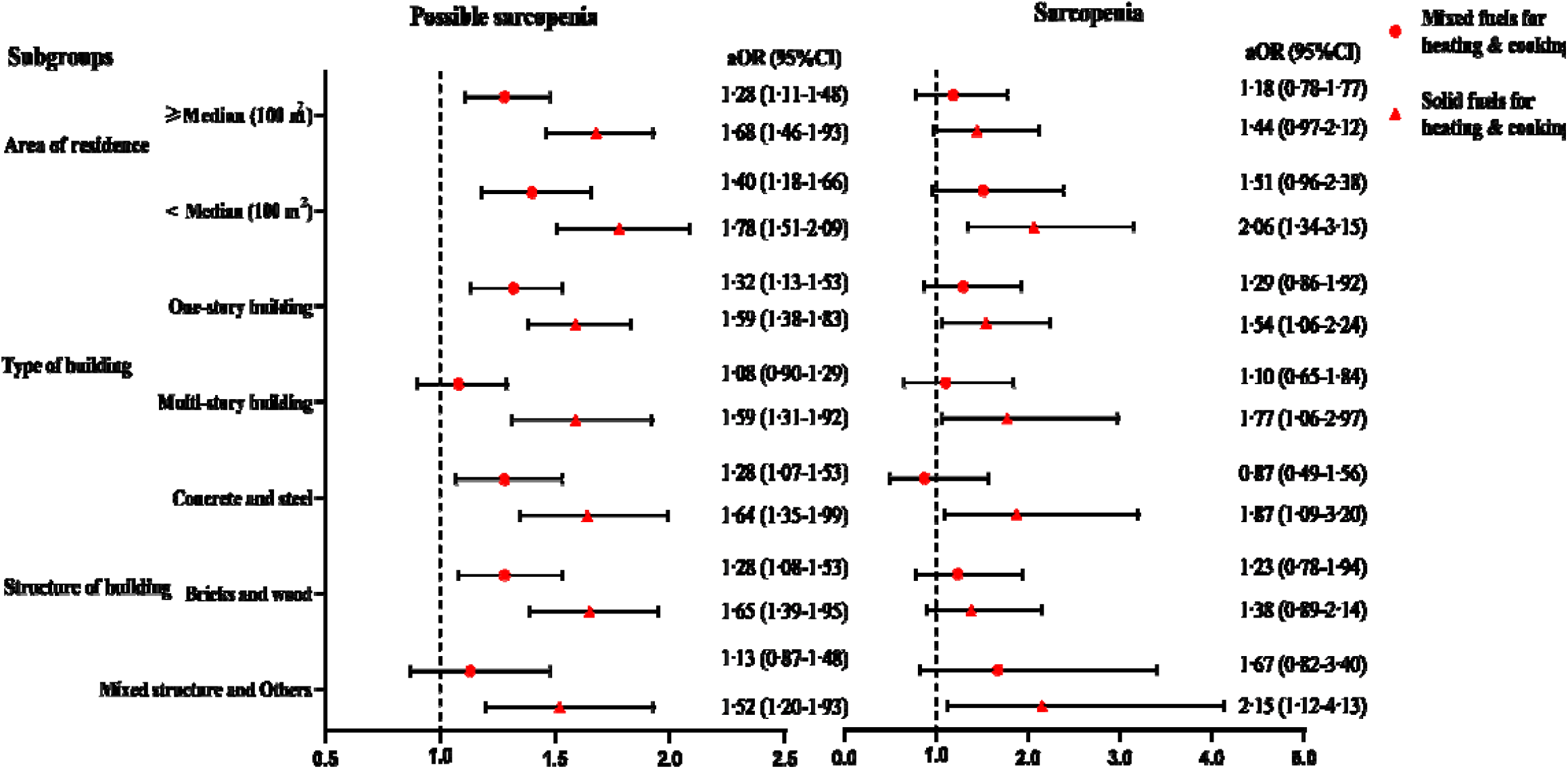
Associations between household solid fuels use and different stages of sarcopenia stratified by residential building characteristics. Note:all the analyses using no sarcopenia as reference. Model adjusted for age, gender, BMI, marriage status, educational level, family’s economic status, cigarette smoking, alcohol consumption, physical activity, and residence.

## References

1 Cruz-Jentoft AJ, Sayer AA. Sarcopenia. Lancet. 2019;393:(10191):2636–46.

2 Mayhew AJ, Amog K, Phillips S,et al The prevalence of sarcopenia in community-dwelling older adults, an exploration of differences between studies and within definitions: a systematic review and meta-analyses. Age Ageing. 2019;48:(1):48–56.

3 Sun MY, Chang CL, Lu CY, Wu SY, Zhang JQ. Sarcopenia as an Independent Risk Factor for Specific Cancers: A Propensity Score-Matched Asian Population-Based Cohort Study. Nutrients. 2022;14:(9).

4 Gao K, Cao LF, Ma WZ,et al Association between sarcopenia and cardiovascular disease among middle-aged and older adults: Findings from the China health and retirement longitudinal study. EClinicalMedicine. 2022;44:101264.

5 Xia L, Zhao R, Wan Q,et al Sarcopenia and adverse health-related outcomes: An umbrella review of meta-analyses of observational studies. Cancer Med. 2020;9:(21):7964–78.

6 Liguori I, Russo G, Aran L,et al Sarcopenia: assessment of disease burden and strategies to improve outcomes. Clin Interv Aging. 2018;13:913–27.

7 Cruz-Jentoft AJ, Bahat G, Bauer J,et al Sarcopenia: revised European consensus on definition and diagnosis. Age Ageing. 2019;48:(1):16–31.

8 Chen LK, Liu LK, Woo J,et al Sarcopenia in Asia: consensus report of the Asian Working Group for Sarcopenia. J Am Med Dir Assoc. 2014;15:(2):95–101.

9 GBD. Global burden of 87 risk factors in 204 countries and territories, 1990-2019: a systematic analysis for the Global Burden of Disease Study 2019. Lancet. 2020;396:(10258):1223–49.

10 Zare SM, Lafontaine A, Yang J,et al Association between Outdoor Air Pollution Exposure and Handgrip Strength: Findings from the French CONSTANCES Study. Environ Health Perspect. 2022;130:(5):57701.

11 Tung NT, Lee YL, Lin SY,et al Associations of ambient air pollution with overnight changes in body composition and sleep-related parameters. Sci Total Environ. 2021;791:148265.

12 Lai Z, Yang Y, Qian ZM, Vaughn MG, Tabet M, Lin H. Is ambient air pollution associated with sarcopenia? Results from a nation-wide cross-sectional study. Age Ageing. 2022;51:(11).

13 Chen LK, Woo J, Assantachai P,et al Asian Working Group for Sarcopenia: 2019 Consensus Update on Sarcopenia Diagnosis and Treatment. J Am Med Dir Assoc. 2020;21:(3):300–07.

14 Shi W, Zhang T, Li Y, Huang Y, Luo L. Association between household air pollution from solid fuel use and risk of chronic diseases and their multimorbidity among Chinese adults. Environ Int. 2022;170:107635.

15 Li C, Zhou Y. Residential environment and depressive symptoms among Chinese middle- and old-aged adults: A longitudinal population-based study. Health Place. 2020;66:102463.

16 Krems C, Lhrmann PM, Neuhuser-Berthold M. Physical activity in young and elderly subjects. J Sports Med Phys Fitness. 2004;44:(1):71–76.

17 Hystad P, Duong M, Brauer M,et al Health Effects of Household Solid Fuel Use: Findings from 11 Countries within the Prospective Urban and Rural Epidemiology Study. Environ Health Perspect. 2019;127:(5):57003.

18 Deng Y, Gao Q, Yang D,et al Association between biomass fuel use and risk of hypertension among Chinese older people: A cohort study. Environ Int. 2020;138:105620.

19 Zhao Y, Hu Y, Smith JP, Strauss J, Yang G. Cohort profile: the China Health and Retirement Longitudinal Study (CHARLS). Int J Epidemiol. 2014;43:(1):61–68.

20 Hu Y, Peng W, Ren R, Wang Y, Wang G. Sarcopenia and mild cognitive impairment among elderly adults: The first longitudinal evidence from CHARLS. J Cachexia Sarcopenia Muscle. 2022;13:(6):2944–52.

21 Wu X, Li X, Xu M, Zhang Z, He L, Li Y. Sarcopenia prevalence and associated factors among older Chinese population: Findings from the China Health and Retirement Longitudinal Study. Plos One. 2021;16:(3):e247617.

22 Wen X, Wang M, Jiang CM, Zhang YM. Anthropometric equation for estimation of appendicular skeletal muscle mass in Chinese adults. Asia Pac J Clin Nutr. 2011;20:(4):551–56.

23 Yang M, Hu X, Wang H, Zhang L, Hao Q, Dong B. Sarcopenia predicts readmission and mortality in elderly patients in acute care wards: a prospective study. J Cachexia Sarcopenia Muscle. 2017;8:(2):251–58.

24 Chen Y, Shen G, Liu W,et al Field measurement and estimate of gaseous and particle pollutant emissions from cooking and space heating processes in rural households, northern China. Atmos Environ. 2016;125:265–71.

25 Chen Z, Ho M, Chau PH. Prevalence, Incidence, and Associated Factors of Possible Sarcopenia in Community-Dwelling Chinese Older Adults: A Population-Based Longitudinal Study. Front Med (Lausanne). 2021;8:769708.

26 Holtermann A, Schnohr P, Nordestgaard BG, Marott JL. The physical activity paradox in cardiovascular disease and all-cause mortality: the contemporary Copenhagen General Population Study with 104 046 adults. Eur Heart J. 2021;42:(15):1499–511.

27 Yu K, Qiu G, Chan KH,et al Association of Solid Fuel Use With Risk of Cardiovascular and All-Cause Mortality in Rural China. JAMA. 2018;319:(13):1351–61.

28 Ali MU, Yu Y, Yousaf B,et al Health impacts of indoor air pollution from household solid fuel on children and women. J Hazard Mater. 2021;416:126127.

29 Li Q, Jiang J, Wang S,et al Impacts of household coal and biomass combustion on indoor and ambient air quality in China: Current status and implication. Sci Total Environ. 2017;576:347–61.

30 Lin H, Guo Y, Ruan Z,et al Association of Indoor and Outdoor Air Pollution With Hand-Grip Strength Among Adults in Six Low- and Middle-Income Countries. J Gerontol A Biol Sci Med Sci. 2020;75:(2):340–47.

31 de Zwart F, Brunekreef B, Timmermans E, Deeg D, Gehring U. Air Pollution and Performance-Based Physical Functioning in Dutch Older Adults. Environ Health Perspect. 2018;126:(1):17009.

32 Sayer AA, Robinson SM, Patel HP, Shavlakadze T, Cooper C, Grounds MD. New horizons in the pathogenesis, diagnosis and management of sarcopenia. Age Ageing. 2013;42:(2):145–50.

33 Byun HM, Panni T, Motta V,et al Effects of airborne pollutants on mitochondrial DNA methylation. Part Fibre Toxicol. 2013;10:18.

34 Parise G, De Lisio M. Mitochondrial theory of aging in human age-related sarcopenia. Interdiscip Top Gerontol. 2010;37:142–56.

35 Sousa-Victor P, Munoz-Canoves P. Regenerative decline of stem cells in sarcopenia. Mol Aspects Med. 2016;50:109–17.

36 Dhillon RJ, Hasni S. Pathogenesis and Management of Sarcopenia. Clin Geriatr Med. 2017;33:(1):17–26.

37 Ochodo C, Ndetei DM, Moturi WN, Otieno JO. External built residential environment characteristics that affect mental health of adults. J Urban Health. 2014;91:(5):908–27.

